# Tattoos and risk of cutaneous melanoma and non-melanoma skin cancer in France

**DOI:** 10.1101/2025.09.25.25336614

**Authors:** Tingting Mo, Marie Zins, Marcel Goldberg, Céline Ribet, Sofiane Kab, Ines Schreiver, Katherina Siewert, Khaled Ezzedine, Joachim Schüz, Milena Foerster

## Abstract

**Background:** With the increasing popularity of decorative tattooing, which entails the intradermal injection of inks that may contain carcinogens, investigating the related potential skin cancer risk is a public health priority.

**Methods:** We used data from the Cancer Risk Attributable with the Body Art of Tattooing (CRABAT) study, nested in the French national cohort Constances (adults aged 18-69 years recruited in 2012–2018). Tattoo exposure was collected in 2020–23. Skin cancers overall, cutaneous melanoma (CM), and non-melanoma skin cancer (NMSC) diagnosed during 2007–21 were retrieved from national health insurance data. As exposure information was collected after possible disease ascertainment, risks of skin cancer with prior tattoo exposure were assessed using (i) Logistic regression and (ii) retrospective cohort analyses using Cox proportional hazards model.

**Results:** Among 111 074 participants, 1789 skin cancers (1.6%) were recorded (693 CM, 1096 NMSC). No association was found between binary tattoo exposure and any skin cancer type. In logistic regression, tattoo body surface >2 hand palms was associated with lower overall skin cancer risk (OR = 0.21, 95% CI: 0.05–0.83; reference no tattoos). This association was not significant in the Cox model, but the suggestive dose–response relationship remained, with HRs of 1.14, 0.60, and 0.26 for tattoo body surface of 0–1, 1–2, and >2 hand palms, respectively.

**Conclusion:** Large tattoo surfaces were tentatively associated with reduced overall skin cancer risk. While these finding merits further research, small case numbers and the retrospectively collected data might have biased the results.

## Introduction

During the past decades, tattooing has become increasingly popular, particularly in young adults, all over the globe. In industrialised countries, up to 40% of individuals aged 30-40 years report having at least one tattoo (1).

Potential cancer risks from tattoos arise due to the presence of known and/or suspected carcinogens in commercially available tattoo inks (2, 3). Among these substances, stemming mostly from contaminated raw materials, are polycyclic aromatic hydrocarbons bound to carbon black pigments, phototoxic primary aromatic amines in bright-coloured inks, and metals, found in inks of all colours (4, 5). During tattooing the inks are injected into the dermis, internalised by phagocytes, and partly transported to the draining lymph nodes (6–8). Although the cancer risk implications of this exposure route are poorly understood, there is no doubt that it entails high local pigment exposure to the skin and lymph nodes, and consequently oncogenesis is possible in both organs (9–12).

To date, four epidemiological studies on tattoo-associated skin tumour risk were published of which one included too few cases to draw conclusions (13). Using data from the early 2000s, a case-control study restricted to tattooed individuals found an increased risk of basal cell carcinoma at the precise body part that was tattooed, especially for green and yellow pigmented tattoos (14). Two more recent case-control studies on non-melanoma skin cancer (NMSC) and cutaneous melanoma (CM) were published this year, using Swedish and US registry data, respectively (15, 16). While tattoo exposure did not alter the risk for NMSC in the Swedish study, in the US study from Utah, a decreased risk for CM in the highest exposure category contrasted by an increased risk for CM in-situ in men with small tattoos was seen.

The Cancer Risk Associated with the Body Art of Tattooing (CRABAT) study, initiated by the International Agency for Research of Cancer, is so far the largest cohort of tattooed individuals with the future potential to investigate cancer risk associated with tattooing, including the risk of CM - the deadliest skin tumor - and NMSC. Although the follow-up time of CRABAT is still too short to provide prospective results, the first results relying on retrospectively collected data on tattoo exposure and skin cancer diagnosed from 2007-2020/21 are presented here.

## Methods

### Study design and population

CRABAT is an ongoing cohort study investigating the potential role of tattoos in the development and progression of cancer (16). The study is nested within the French national cohort Consultants des Centres d’Examens de Santé (Constances). Baseline recruitment of Constances took place from 2012 to 2019, during which 220000 voluntary participants aged 18-69 at inclusion were sampled to represent the country’s age, sex, and socioeconomic structure (17). Constances provides a unique research infrastructure for epidemiological research with a wealth of self-reported data on sociodemographic, lifestyle, and medical factors collected during baseline and annual follow-up questionnaires. Objective data on health outcomes is ensured through record linkage of cohort members to national health databases (Système National des Données de Santé; SNDS). These databases log information on all medical services delivered by hospitals, private practices, and pharmacies that are reimbursed by the national public health insurance, with almost full population coverage.

CRABAT collected tattoo exposure within Constances first in the annual follow-up questionnaire 2020 via the screening question “Are you tattooed?”, then the follow-up question about tattooed surface larger than 1 hand surface or not. In the second phase, the validated Epidemiological Tattoo Assessment Tool (EpiTAT questionnaire) was sent to the tattooed subpopulation from July to December 2023 for more detailed exposure information including precise tattoo body surface, colour, time period, tattoo sun exposure etc. (18). In February 2024, the collected exposure data were merged with prior sociodemographic, lifestyle, and medical data for both tattooed and non-tattooed participants who responded to the 2020 follow-up.

Based on CRABAT, this study included participants who provided tattoo information on 2020. We further excluded participants with missing data on exposure or outcome, permanent make-up/facial tattoos, tattoo removal, and participants of very dark skin (phototype VI) or with albinism as they have very low or very high skin cancer risk compared to other skin types, respectively (19) (Figure 1).

**Figure 1.**
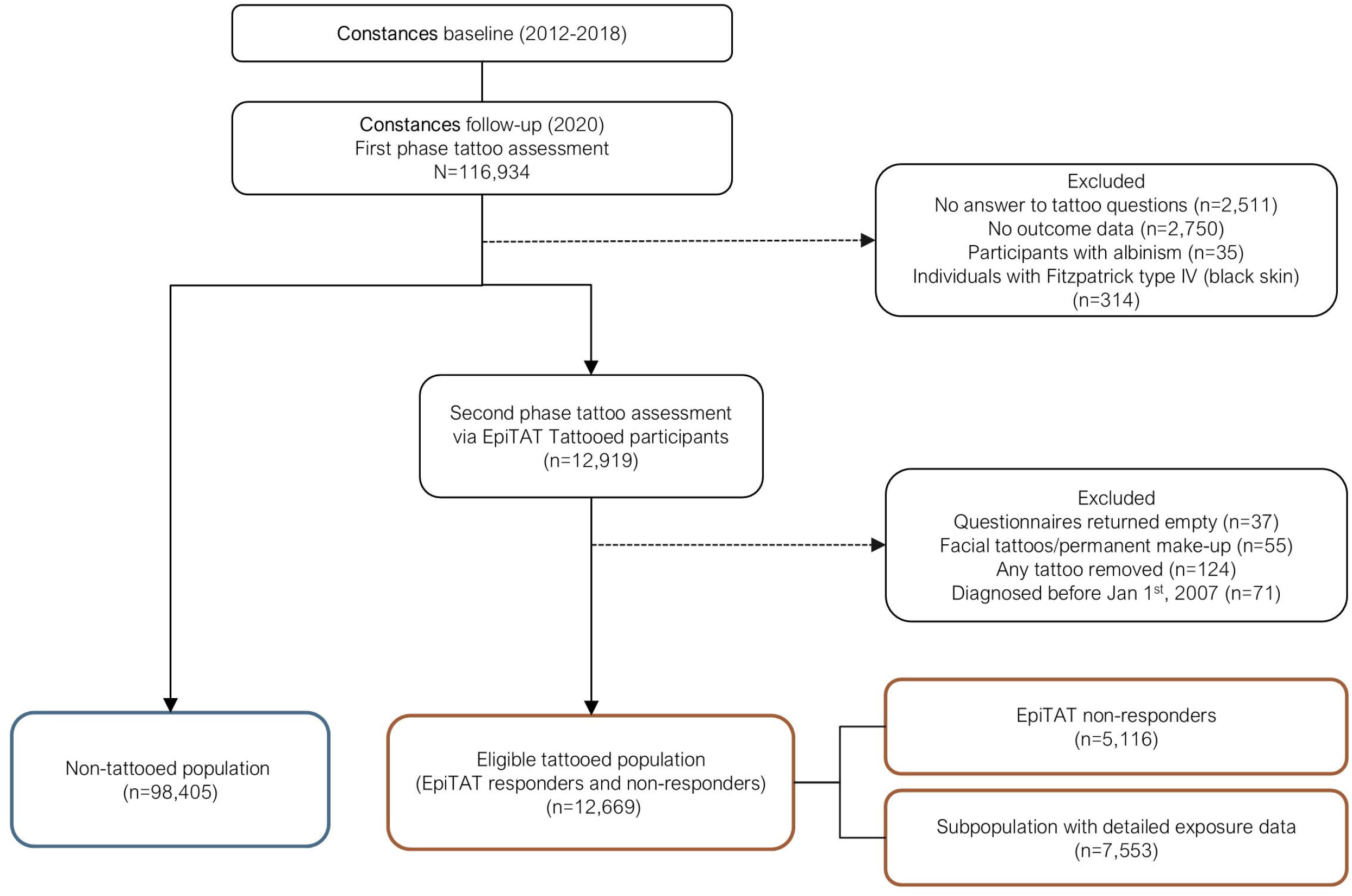
Flow chart of determining the study population from the Cancer Risk Attributed to the Body Art of Tattooing (CRABAT) study.

### Exposure data

Binary tattoo status (yes, no) was retrieved from the Constances follow-up questionnaire 2020/21. Tattooed body surface (TBS) measured in hand surfaces was derived as composite variable from the same questionnaire and the detailed exposure assessment in 2023. TBS was categorised as non-tattooed, 0-1 (participants reporting TBS≤1 in 2020/21 and TBS≤1 or non-response in 2023), >1-2 (TBS reported in 2023), and >2 (TBS reported in 2023). Participants who reported a tattooed area of >1 and did not respond in 2023 were excluded from dose-dependent models due to potential exposure misclassification. For participants responding TBS<1 in 2020/21 but not responding in 2023, tattoo surface was set to 0.5 hand surfaces (to be included in dose-dependent models on “incremental tattoo surface”). To account for potential interaction of sunlight, tattoo sun exposure was categorised as non-tattooed, tattooed not sun-exposed, or tattooed sun-exposed.

First tattoo date was assessed through multiple choice categories and was set as the midpoint of the indicated time period (last year, >1-5 years ago, >5-10 years ago, >10-15 years ago, >15 years ago), to 20 years ago if “>15 years ago” was answered, and earliest at age 16. To minimise selection bias, we imputed the first tattoo date in the 40% of tattooed participants that did not answer the EpiTAT question in 2023, allowing to include this participant group in time-to-event models (multiple imputation described in Supplementary Methods S1, and Table S1-S2).

### Outcome data

Data on cases of CM (ICD-10 code C43) and NMSC (C44) treated in public and/or private hospitals, and their respective hospital entry date, were retrieved from the SNDS database for the time period 1^st^ Jan 2007 to 2020/21 (date of first exposure assessment). Data on tumour location, subtypes, or grade were not available. Those participants whose date of first tattoo could be determined and who developed a CM or NMSC before that date were treated as non-tattooed (n=2).

### Additional self-reported covariate data retrieved from Constances

Data on sex, age, highest education (no diploma, high school degree, bachelor degree, master degree and higher; according to French education system), household disposable income (≤2100€, 2100 to <2800€, 2800 to <4200€, ≥4200€), smoking status (never smoked, former smoker, current smoker), and body mass index (BMI) were retrieved from the Constances baseline questionnaires. Because barriers to health care might lead to differential underdiagnosis of skin tumours if more/less common in the exposed compared to the non-exposed population, a composite variable (no barriers; barriers at least reports once) was created from recurrently asked questions on financial, spatial and organisational barriers to healthcare. Dermal skin cancer risk factors including frequency of sunburns during adolescence (never, 1–5 times; 6–10 times; every summer; I don’t know), maximum intensity of tan (absent; light; bright; dark; very dark), and skin type assessed according to the Fitzpatrick scale (very fair [white]; very fair and may be tanned; moderately fair; matt; dark brown) were collected in a dedicated skin questionnaire during the 2018 Constances follow-up (20).

### Statistical analysis

As exposure data were obtained after potential disease ascertainment, we used two approaches:

i. Logistic regression to assess risks of historical skin cancer in relation to prior tattoo exposure (71 observations excluded because of first tattoo in the year of or after diagnosis). Exposure variables were tattooed (yes, no), TBS (not tattooed, 0-1, >1-2, and >2 hand palms), and within the tattooed population: incremental tattoo surface, and tattoo sun exposure (not sun-exposed, and sun-exposed). Two levels of adjustment were performed adjusting variables at Constances’s baseline: model 1 adjusted for sex, age, education level, smoking status, and BMI; model 2 additionally adjusted for household disposable income, skin type, maximum intensity of tan, frequency of sunburn during adolescence, and barriers to healthcare (covariate selection described in Supplementary Methods S2, Table S3, and Figure S1). Missing values in covariates were coded as a separate category. Moreover, we fitted model 2 with and without TBS exposure to explore independency of the tattoo exposure from other covariates.
ii. Survival analysis using Cox proportional hazards regression assuming no exposure misclassification or differential losses in that population group. Exposure variables and covariates remained unchanged from logistic regression (except for age, because models were fitted on the age-time scale). Two alternative entry dates were considered, January 1^st^, 2007 (i.e. date from which on SNDS data was available) or the individual Constance baseline date. To account for potential birth cohort effect linked to the marked changes in sunscreen use during childhood and adult tattoo prevalence that occurred from people born before 1980 to thereafter (*note: as those changes occurred gradually, the cut-off year can only be an approximation*), analyses were stratified by age in 2020 (<40, ≥40). Considering being tattooed as time-varying exposure, the dataset was split (STATA: stsplit) on the date of first tattoo; thus tattooed participants will contribute person-years to the exposed group only from that date onwards, being before counted as non-exposed.

As sensitivity analyses, we repeated the logistic regression and survival metrics restricted to participants aged over 40 at exposure assessment, and the survival metrics among those who provided the first tattoo date.

## Results

Of 111074 eligible participants, 12669 (11.5%) reported tattooed in the 2020 questionnaire (Table 1, Figure 1). Full exposure information was available for 60% of participants (7553/12669), and tattoo surface could be determined for >80% of participants, about half of whom reported a tattooed surface of 0-1 hand palm. Women were tattooed more often compared with men though men more often had larger tattoos (>2 hand palms). Tattooed participants were younger, particularly among those with large tattooed body surfaces. Tattooed participants had a lower education level and household disposable incomes, and more often reported barriers to seeking health care. Also, tattooed participants were more likely to have darker skin and more often had sunburns before adulthood, particularly among those with large tattooed surfaces (Table 2). Besides of the skin characteristics selected for analyses, tattooed participants tended to have more moles, freckles, and sunburns during adulthood (Table S4).

**Table 1:** Sociodemographic characteristics in 111,074 eligible participants of the Cancer Risk Attributed to the Body art of Tattooing (CRABAT) study, for tattooed and non-tattooed individuals, and for tattooed individuals by tattooed body surface.

| Characteristic | Tattoo status, 2020 Constances respondents |  | Tattoo size, among all tattooed<sup>a</sup> |  |  |  |
| --- | --- | --- | --- | --- | --- | --- |
|  | Tattooed | Non-tattooed | 0–1 hand surface | 1–2 hand surfaces | >2 hand surfaces | Size missing |
|  | N (% among full sample, in category) |  | N (% among the tattooed categories) |  |  |  |
| No. of participants | 12669 (11.5%) | 98405 (89.5%) | 6712 (53.3%) | 2001 (15.9%) | 1633 (13.0%) | 2323 (18.5%) |
| Sex, N (%) |  |  |  |  |  |  |
| Male | 4736 (37.4%) | 45606 (46.3%) | 2356 (35.1%) | 706 (35.3%) | 682 (41.8%) | 992 (42.7%) |
| Female | 7933 (62.6%) | 52799 (53.7%) | 4356 (64.9%) | 1295 (64.7%) | 951 (58.2%) | 1331 (57.3%) |
| Age at baseline, median (IQR) | 40 (32, 49) | 51 (40, 61) | 42 (34, 52) | 38 (31, 46) | 37 (30, 45) | 37 (30, 45) |
| BMI, kg/m<sup>2</sup>, N (%) known<sup>b</sup> | 12531 (98.9%) | 97122 (98.7%) | 6632 (98.8%) | 1985 (99.2%) | 1619 (99.1%) | 2295 (98.8%) |
| Median (IQR) | 24.1 (21.6, 27.2) | 24.2 (21.8, 27.0) | 23.9 (21.5, 27.0) | 24.0 (21.6, 26.9) | 24.4 (21.9, 27.6) | 24.4 (21.9, 27.7) |
| Smoking status, N (%) known<sup>b</sup> | 12517 (98.8%) | 97061 (98.6%) | 6628 (98.7%) | 1983 (99.1%) | 1619 (99.1%) | 2287 (98.5%) |
| Never | 4296 (34.3%) | 49340 (50.8%) | 2275 (34.3%) | 721 (36.4%) | 593 (36.6%) | 707 (30.9%) |
| Former | 4911 (39.2%) | 35753 (36.8%) | 2695 (40.7%) | 759 (38.3%) | 606 (37.4%) | 851 (37.2%) |
| Current | 3310 (26.4%) | 11968 (12.3%) | 1658 (25.0%) | 503 (25.4%) | 420 (25.9%) | 729 (31.9%) |
| Education, N (%) known<sup>b,c</sup> | 12459 (98.3%) | 96596 (98.2%) | 6604 (98.4%) | 1974 (98.7%) | 1612 (98.7%) | 2269 (97.7%) |
| No diploma | 3104 (24.9%) | 19403 (20.1%) | 1659 (25.1%) | 412 (20.9%) | 385 (23.9%) | 648 (28.6%) |
| High school degree | 2642 (21.2%) | 14047 (14.5%) | 1218 (18.4%) | 456 (23.1%) | 361 (22.4%) | 607 (26.8%) |
| Bachelor degree | 4630 (37.2%) | 35272 (36.5%) | 2487 (37.7%) | 797 (40.4%) | 604 (37.5%) | 742 (32.7%) |
| Master degree and higher | 2083 (16.7%) | 27874 (28.9%) | 1240 (18.8%) | 309 (15.7%) | 262 (16.3%) | 272 (12.0%) |
| Household disposable income, N (%) known<sup>b</sup> | 11797 (93.1%) | 92188 (93.7%) | 6268 (93.4%) | 1869 (93.4%) | 1534 (93.9%) | 2126 (91.5%) |
| ≤2100€ | 3096 (26.2%) | 15104 (16.4%) | 1504 (24.0%) | 455 (24.3%) | 448 (29.2%) | 689 (32.4%) |
| 2100 to <2800€ | 2045 (17.3%) | 13370 (14.5%) | 1047 (16.7%) | 324 (17.3%) | 298 (19.4%) | 376 (17.7%) |
| 2800 to <4200€ | 4294 (36.4%) | 30374 (32.9%) | 2301 (36.7%) | 731 (39.1%) | 512 (33.4%) | 750 (35.3%) |
| ≥4200€ | 2362 (20.0%) | 33340 (36.2%) | 1416 (22.6%) | 359 (19.2%) | 276 (18.0%) | 311 (14.6%) |
| Barriers to healthcare<sup>d</sup> |  |  |  |  |  |  |
| No barriers | 9701 (76.6%) | 83316 (84.7%) | 5259 (78.4%) | 1500 (75.0%) | 1195 (73.2%) | 1747 (75.2%) |
| Barriers reported at least once | 2968 (23.4%) | 15089 (15.3%) | 1453 (21.6%) | 501 (25.0%) | 438 (26.8%) | 576 (24.8%) |

<sup>a</sup> Tattoo surface retrieved from second phase exposure assessment in 2023 (EpiTAT exposure questionnaire). In addition, participants who only took part in the first phase exposure assessment in 2020 (Constances follow-up questionnaire) and who therein reported tattoo surface of “≤ 1 hand surface” (vs “>1 hand surface”) were added to the lowest exposure category.
<sup>b</sup> N (%) known indicates the number and percentage of non-missing values for each variable.
<sup>c</sup> Original categorisation according to French education system, i.e. “high school degree” equals the French “baccalauréat général” after 12 years of schooling, “bachelor degree” equals a university degree obtained after up to four years (“bac +2/3/4”), and “master degree” equals a university degree obtained after at least five years (“bac +5 et supérieur”).
<sup>d</sup> Dichotomous variable composed from three variables enquiring whether in the last 12 months the participant or a family member had to forego healthcare due to (1) financial problems, (2) long distance to medical services or (3) too long waiting times for an appointment. Variable was coded “yes” if any of these were answered with “yes” in at least one of (up to) six follow-up questionnaires.

**Table 2.** Skin characteristics of the CRABAT participants, for tattooed and non–tattooed individuals, and for tattooed individuals by tattooed body surface.

| Characteristic | Tattoo status, 2020 Constances respondents |  | Tattoo size, among all tattooed<sup>a</sup> |  |  |  |
| --- | --- | --- | --- | --- | --- | --- |
|  | Tattooed | Not tattooed | 0–1 hand surface | 1–2 hand surface | >2 hand surfaces | Tattooed, size missing |
|  | N (% among full sample, in category) |  | N (% among the tattooed categories) |  |  |  |
| No of participants (% row) | 12669 (11.5%) | 98405 (89.5%) | 6712 (53.3%) | 2001 (15.9%) | 1633 (13.0%) | 2323 (18.5%) |
| Maximum intensity of tan, N (%) known<sup>b</sup> | 7327 (57.8%) | 63627 (64.7%) | 3991 (59.5%) | 1226 (61.3%) | 984 (60.3%) | 1126 (48.5%) |
| Absent | 72 (1.0%) | 988 (1.6%) | 42 (1.1%) | 8 (0.7%) | 15 (1.5%) | 7 (0.6%) |
| Light | 1021 (13.9%) | 12062 (19.0%) | 536 (13.4%) | 154 (12.6%) | 162 (16.5%) | 169 (15.0%) |
| Bright | 2010 (27.4%) | 19685 (30.9%) | 1135 (28.4%) | 351 (28.6%) | 250 (25.4%) | 274 (24.3%) |
| Dark | 3852 (52.6%) | 28995 (45.6%) | 2084 (52.2%) | 653 (53.3%) | 499 (50.7%) | 616 (54.7%) |
| Very dark | 372 (5.1%) | 1897 (3.0%) | 194 (4.9%) | 60 (4.9%) | 58 (5.9%) | 60 (5.3%) |
| Skin type, N (%) known<sup>b,c</sup> | 7266 (57.4%) | 62944 (64.0%) | 3956 (58.9%) | 1225 (61.2%) | 975 (59.7%) | 1110 (47.8%) |
| Very fair, white | 212 (2.9%) | 1660 (2.6%) | 104 (2.6%) | 30 (2.4%) | 30 (3.1%) | 48 (4.3%) |
| Very fair, and may be tanned | 1230 (16.9%) | 9479 (15.1%) | 644 (16.3%) | 220 (18.0%) | 191 (19.6%) | 175 (15.8%) |
| Moderately fair | 3837 (52.8%) | 34339 (54.6%) | 2110 (53.3%) | 649 (53.0%) | 495 (50.8%) | 583 (52.5%) |
| Matt | 1842 (25.4%) | 16500 (26.2%) | 1018 (25.7%) | 303 (24.7%) | 241 (24.7%) | 280 (25.2%) |
| Dark brown | 145 (2.0%) | 966 (1.5%) | 80 (2.0%) | 23 (1.9%) | 18 (1.8%) | 24 (2.2%) |
| Sunburn during adolescence, N (%) known<sup>b</sup> | 7313 (57.7%) | 63303 (64.3%) | 3987 (59.4%) | 1231 (61.5%) | 972 (59.5%) | 1123 (48.3%) |
| Never | 758 (10.4%) | 6783 (10.7%) | 400 (10.0%) | 124 (10.1%) | 92 (9.5%) | 142 (12.6%) |
| 1–5 times | 1988 (27.2%) | 17572 (27.8%) | 1072 (26.9%) | 341 (27.7%) | 270 (27.8%) | 305 (27.2%) |
| 6–10 times | 1548 (21.2%) | 13797 (21.8%) | 872 (21.9%) | 256 (20.8%) | 199 (20.5%) | 221 (19.7%) |
| Every summer | 2273 (31.1%) | 18440 (29.1%) | 1229 (30.8%) | 389 (31.6%) | 312 (32.1%) | 343 (30.5%) |
| I don't know | 746 (10.2%) | 6711 (10.6%) | 414 (10.4%) | 121 (9.8%) | 99 (10.2%) | 112 (10.0%) |

<sup>a</sup> Tattoo surface retrieved from second phase exposure assessment in 2023 (EpiTAT exposure questionnaire). In addition, participants who only took part in the first phase exposure assessment in 2020 (Constances follow-up questionnaire) and who therein reported tattoo surface of “≤ 1 hand surface” (vs “>1 hand surface”) were added to the lowest exposure category.
<sup>b</sup> N (%) known indicates the number and percentage of non-missing values for each variable.
<sup>c</sup> Categorization according to the Fitzpatrick scale. Individuals with Fitzpatrick type IV (black skin, n=292) were excluded from the study because of their very low skin cancer risk.

Exploring selection bias, sociodemographic and skin characteristics were compared between tattooed participants who answered to the EpiTAT questionnaire in 2023 and those who did not. Compared with EpiTAT non-responders, responders were more likely to be female, have lower BMI, not currently smoking, have higher education and household disposable income (Table S5). Additionally, EpiTAT responders were more likely to have more moles and lighter skin and more sunburns (Tables S6).

Overall, 1789 skin cancer cases (693 CM, 1096 NMSC) were recorded, 15-year incidence percentages were 2-3 times higher in non-tattooed compared with tattooed participants (Table 3). In logistic regression models, after full adjustment, being tattooed was not associated with overall skin cancer risk (Table 3). However, compared with participants without tattoo, there was a significant risk decrease of 79% in the highest exposure category of TBS >2 hand surface (OR=0.21, 95% CI=0.05 to 0.83). The same pattern was seen for NMSC, though without reaching statistical significance (OR=0.39, 95% CI=0.10 to 1.58), whilst no CM cases occurred in the highest tattoo exposure category. In the tattooed population, ORs were consistently elevated for overall skin cancer, CM and NMSC (OR=1.75, 2.81, and 1.17, respectively) if tattoos were not exposed to sunlight (compared to exposed), but case numbers were small and estimates lacked precision. Comparison of multivariate models with and without TBS supported the independency of the tattoo exposure from other covariates (Table S7).

**Table 3.** Odd’s ratios (ORs) and confidence intervals (CIs) for the cross–sectional relationship of different tattoo exposure variables with overall skin cancer, melanoma, and non–melanoma in the Cancer Risk Attributable to the Body Art of Tattooing (CRABAT) study.

| Exposure variable | n/category<sup>a</sup> | Overall skin cancer |  | Cutaneous melanoma |  | Non-melanoma skin cancer |  |
| --- | --- | --- | --- | --- | --- | --- | --- |
|  |  | Cases (% of category) | OR (95% CI)<sup>b</sup> | Cases (% of category) | OR (95% CI)<sup>b</sup> | Cases (% of category) | OR (95% CI)<sup>b</sup> |
| **Tattooed** |  |  |  |  |  |  |  |
| N Total | 111074 | 1789 (1.6%) |  | 693 (0.6%) |  | 1096 (1.0%) |  |
| No | 98405 | 1702 (1.8%) | ref | 659 (0.7%) | ref | 1043 (1.1%) | ref |
| Yes | 12669 | 87 (0.7%) | 0.92 (0.74-1.15) | 34 (0.3%) | 0.80 (0.56-1.14) | 53 (0.4%) | 1.02 (0.76-1.35) |
| **Tattooed body surface** |  |  |  |  |  |  |  |
| N Total | 108751 | 1771 (1.6%) |  | 687 (0.6%) |  | 1096 (1.0%) |  |
| Not tattooed | 98405 | 1702 (1.8%) | ref | 659 (0.7%) | ref | 1043 (1.1%) | ref |
| 0-1 hand palm | 6712 | 61 (0.9%) | 1.04 (0.80-1.35) | 26 (0.4%) | 1.01 (0.68-1.51) | 35 (0.5%) | 1.06 (0.75-1.49) |
| 1-2 hand palms | 2001 | 6 (0.3%) | 0.47 (0.21-1.05) | 2 (0.1%) | 0.33 (0.08-1.35) | 4 (0.2%) | 0.58 (0.22-1.55) |
| >2 hand palms | 1633 | 2 (0.1%) | 0.21 (0.05-0.83) | No obs | No obs | 2 (0.1%) | 0.39 (0.10-1.58) |
| **Only amongst the tattooed :** |  |  |  |  |  |  |  |
| Incremental among tattooed | 10346 | 69 (0.7%) | 0.85 (0.71-1.02) | 28 (0.3%) | 0.54 (0.28-1.03) | 41 (0.4%) | 0.92 (0.79-1.08) |
| **Sun exposure of tattoo** |  |  |  |  |  |  |  |
| N Total | 5978 | 26 (0.4%) |  | 12 (0.2%) |  | 14 (0.2%) |  |
| Not sun exposed | 1034 | 9 (0.9%) | 1.75 (0.71-4.32) | 4 (0.4%) | 2.81 (0.70-11.25) | 5 (0.5%) | 1.17 (0.31-4.41) |
| Sun exposed | 4944 | 17 (0.3%) | ref | 8 (0.2%) | ref | 9 (0.2%) | ref |

<sup>a</sup> Due to the different numbers of missing values for each tattoo-related variable, the number of participants were slightly different.
<sup>b</sup> Model adjusted for biological sex, age at Constance's baseline, education level, smoking status, BMI, household disposable income, skin type, maximum intensity of tan, frequency of sunburn during adolescence, and barriers to healthcare.

In the main Cox regression model (with imputed date of first tattoo for ~40% and study entry Jan 1^st^, 2007), the results were comparable though lacked precision: Binary tattoo exposure was not related to any type of skin cancer, but a non-significant risk decrease of 74% was seen with TBS of >2 hand surface (HR=0.26, 95% CI=0.07 to 1.05) (Table 4).

**Table 4.** Hazard ratios (HRs) and confidence intervals (CIs) of Cox proportional hazard regression for the association of different tattoo exposure and risk of overall skin cancer, melanoma, and non-melanoma, in the whole population with study entry date on 1 Jan, 2007 (start of outcome assessment via French health database). Tattoo exposure was considered as time-varying exposure, meaning that an individual contributes person-years to the non-tattooed group until the date of first tattoo, when the individual changes exposure status and will be contributing to the exposed population thereafter. Date of first tattoo was imputed in ~40% of tattooed participants, i.e. those with only minimal exposure data (non-respondents to EpiTAT).

| Imputed in 10% of tattooed participants, ref: those with only minimal exposure data (non-respondents to ZPT111). |  |  |  |  |  |  |  |
| --- | --- | --- | --- | --- | --- | --- | --- |
| Exposure | n/category<sup>a</sup> | Overall skin |  | Melanoma |  | Non-melanoma |  |
|  |  | Cases (% of category) | HR (95% CI)<sup>b</sup> | Cases (% of category) | HR (95% CI)<sup>b</sup> | Cases (% of category) | HR (95% CI)<sup>b</sup> |
| **Tattooed** |  |  |  |  |  |  |  |
| N Total | 111074 | 1789 (1.6%) |  | 693 (0.6%) |  | 1096 (1.0%) |  |
| No | 98405 | 1702 (1.7%) | ref | 659 (0.7%) | ref | 1043 (1.1%) | ref |
| Yes | 12669 | 87 (0.7%) | 1.03 (0.82-1.30) | 34 (0.3%) | 0.96 (0.66-1.38) | 53 (0.4%) | 1.08 (0.80-1.46) |
| **Tattooed body surface** |  |  |  |  |  |  |  |
| N Total | 108751 | 1771 (1.6%) |  | 687 (0.6%) |  | 1084 (1.0%) |  |
| Not tattooed | 98405 | 1702 (1.7%) | ref | 659 (0.7%) | ref | 1043 (1.1%) | ref |
| 0-1 hand palm | 6712 | 61 (0.9%) | 1.14 (0.87-1.49) | 26 (0.4%) | 1.19 (0.79-1.79) | 35 (0.5%) | 1.09 (0.76-1.57) |
| 1-2 hand palms | 2001 | 6 (0.3%) | 0.60 (0.27-1.34) | 2 (0.1%) | 0.46 (0.11-1.86) | 4 (0.2%) | 0.71 (0.26-1.89) |
| >2 hand palms | 1633 | 2 (0.1%) | 0.26 (0.07-1.05) | No obs | No obs | 2 (0.1%) | 0.47 (0.12-1.90) |
| **Only amongst the tattooed :** |  |  |  |  |  |  |  |
| Incremental among tattooed | 10346 | 69 (0.7%) | 0.91 (0.80-1.03) | 28 (0.3%) | 0.75 (0.52-1.06) | 41 (0.4%) | 0.96 (0.86-1.07) |
| **Sun exposure of tattoo** |  |  |  |  |  |  |  |
| N Total | 5978 | 26 (0.4%) |  | 12 (0.2%) |  | 14 (0.2%) |  |
| Not sun exposed | 1034 | 9 (0.9%) | 1.84 (0.75-4.51) | 4 (0.4%) | 2.90 (0.75-11.14) | 5 (0.5%) | 1.30 (0.35-4.84) |
| Sun exposed | 4944 | 17 (0.3%) | ref | 8 (0.2%) | ref | 9 (0.2%) | ref |

<sup>a</sup> Due to the different numbers of missing values for each tattoo-related variable, the number of participants were slightly different.
<sup>b</sup> Considering tattooed or not as time-varying, the dataset was split on date of first tattoo. Cox regression models were performed based on an age-time scale, with stratification of age at first exposure assessment in 2020/21 (<40, ≥40) and adjustment for biological sex, education level, smoking status, BMI, household disposable income, barriers to health care, maximum intensity of tan, skin type, frequency of sunburns during adolescence.

### Sensitivity analyses

When time 0 was set to the baseline of Constances (excluding 854 cases that were diagnosed before and thus substantially reducing power), the association was slightly attenuated (Table S8). Similarly, restricting analyses to participants over age 40 at first exposure assessment to avoid exposure misclassification only led to minimal attenuation of cross-sectional (OR=0.24, 95% CI=0.06 to 0.96) and longitudinal associations (HR=0.30, 95% CI=0.07 to 1.19), and the suggestive trend of larger TBS with lower risk of skin cancer remained (Table S9-S10). In survival analyses restricted to participants with known first tattoo date (i.e. excluding participants with imputed dates), results were comparable but hampered by even fewer outcomes, particularly in higher exposure categories (Tables S11).

## Discussion

Using the data of the CRABAT study on tattoo-associated long-term health effects, our results present the first estimates on the tattoo-related risk for CM and NMSC on a large scale. As a strength, the analyses was based on detailed data on tattoo exposure assessed via a validated questionnaire (18) and potential confounders. While binary tattoo exposure was not associated with neither skin cancer type, there were some indications for dose-dependent decreases in risk for CM and NMSC with increasing tattooed body surface. For example, the risk of overall skin cancer reduced 79% in the logistic regression model for the highest exposure category. However, the same association failed to reach statistical significance when analysing the data using a retrospective cohort approach. More general, the results are strongly limited by small case numbers that led to often imprecise effect estimates, particularly in the higher exposure categories, and by the fact that full exposure information was only available for ~60% of tattooed participants.

The suggestive dose dependent risk decrease in overall skin cancer, NMSC and CM (although no cases in highest exposure category) are in line with a recent case-control study of McCarty et al (21), in which a similar dose-dependent risk decrease for CM was observed in the highest tattoo exposure category. Finally, the hypothesis of tattoos increasing skin cancer risk is frequently challenged by publications claiming that the number of published case-studies of dermal neoplasia on tattooed skin seems rather small compared to today’s tattoo popularity (22).

One plausible explanation for decreased skin cancer risk, is that tattoos could protect against ultraviolet radiation (UVR) from sun-light, which is the most important preventable risk factor for CM and NMSC (22, 23). Particularly if dark, and on sun-exposed body parts, tattoos could absorb and prevent backscattering of incoming UVR at the collagen layer, resulting in UVR exposure reduction. When exploring this hypothesis in our data, the direction of the effect pointed in that direction, but the analysis was strongly hampered by small case numbers. Still, onset of UVR-induced squamous cell carcinoma was delayed in black-tattooed compared with sham-tattooed (tattooed without ink) mice irradiated with standard erythema doses of UVR (24). It seems unlikely that the sun-exposed skin proportion covered by tattoos is substantial for vast majority of - even heavily tattooed-people, therefore, the protective effect would be, if anything, very small. Moreover, CM is often attributable to sun exposure during childhood and any UVR-protective effect of tattooing would occur in adulthood, further limiting the likelihood of tattoos as major protective factor against UVR (23). However, the observed declining CM rates in young persons are at least not suggestive of the opposite (i.e. a large increase in CM risk amongst tattooed individuals) (25).

If these findings find replication in prospective studies, additional immunological effects of the tattooing process may be hypothesized as the skin is an exceptionally immunogenic exposure route (which e.g. intradermal smallpox vaccination takes advantage of) (26–28). One possible hypothesis could be T cell priming against melanoma antigens, that could be locally released from tumour cells (precursor) during the tattooing process triggering profound tissue injury and skin cell death (29, 30). These free antigens could be taken up by dendritic cells (DCs), that mature, and migrate to the draining lymph nodes, there subsequent (tumour) antigen-specific naive T cell priming could take place (8, 31, 32). Similar to what happens during vaccination, the activated T cells could increase systemic and local immunosurveillance against CM cells. And because T-cell memory responses would increase and refresh with each new tattoo, protective effects against skin cancer would likely be more pronounced in individuals with larger tattooed surface.

### Limitations of the study

While our results are interesting and merit further research, several circumstances could have substantially biased the results. First, full exposure data were only available for 60% of the tattooed subgroup that revealed potential selection bias as skin cancer rates were higher in tattooed non-responders compared to responders. To account for this circumstance in Cox-regression, we imputed date of first tattoo in the 40% of tattooed participants without full exposure information. And indeed, when fitting Cox-regression in-compared to excluding these participants, the HR for binary tattoo exposure and overall skin cancer jumped from 1.04 to 0.81. This bias could be partly owed to the retrospective study design: in the present analysis tattoo exposure was self-reported in 2020 or 2023, while cancer outcomes occurred between 2007 and 2021. If response behaviour to our detailed questionnaire on tattoo exposure differed in participants with and without skin cancer, recall bias could have occurred that needs to be considered when interpreting these results. However, at least exposure misclassification of tattoo status is unlikely, because minimal exposure information was available for >98% of participants who answered to the Constances follow-up in 2020.

Moreover, some uncertainty remains regarding the temporality of exposure and outcome, as first tattoo date was estimated based on being tattooed in a certain time period (instead of an actual date). However, in participants with reported timing since tattoo, we detected only five cases there temporality between exposure outcome was uncertain, reflecting that first tattoos are generally acquired in early adulthood limiting outcome misclassifications in the remaining 40% of tattooed participants (which would moreover lead, if anything, to underestimation of the protective effect).

In our study, outcome data was obtained from patient records of hospitals. This might have led to outcome misclassification through undetected in-situ or early-stage skin tumours that also be treated in dermatological private practices. In contrast, misclassification for more severe outcomes is less likely as most invasive CM will be treated in hospitals, as well as the more aggressive NMSC subtypes. (Therefore, we assume, despite the lack of data on NMSC subtype, that most included cases will be squamous cell carcinoma.)

Also, we could not retrieve data on tumour locations, diagnostic stage or CM/NMSC subtypes. Tumour locations would be useful, because we would expect tattoo-induced skin tumours to be locally confined to tattooed skin, thus matching both in the analysis design would have been useful. Moreover, without this information, the potential protective effects of tattoos against UVR, that would similarly only decrease risk on tattooed, and sun exposed body parts is difficult to assess.

Selective outcome misclassification might have also occurred because of lack of diagnostic stage. As tattooed participants more often reported barriers to health care, it seems plausible that skin tumours could be diagnosed at later stage on average in that group; also, tumours might be more difficult to detect on tattooed skin that could also lead to late-stage diagnosis. While this would only slightly affect the logistic regression estimates, hazard ratios could be stronger as follow-up times would be artificially prolonged in tattooed participants.

While vast data on confounders was obtained through the Constances data pool, data on sunscreen and tanning bed use was not available. In the above-mentioned case-control study from Utah, sunscreen use seemed to be inversely, and frequency of tanning bed use was strongly, positively correlated with number of tattoo sessions (33). While comparability of the French and US population might not be perfect, we would expect that lack of these factors should not lead to an overestimation of the protective effect. If anything, the opposite could be the case.

Finally, compared to Western cultures, tattoo culture is less popular in many other world regions that as well differ in skin cancer incidence rates. However, we did not adjust for region of birth, because in Constances, only 5% of participants were born outside Europe (data not shown) and because global differences in skin cancer rates are mainly driven by natural skin pigmentation, which we accounted for via adjusting for the Fitzpatrick skin type.

## Conclusions

Given the current debate and concerns about the safety of tattooing, our results do not support the claim that tattoos may cause skin cancer; on the contrary, we observed some indications for an inverse association, especially when tattooed body areas are large which is in line with a recent case-control study on the same topic (3, 34). As case numbers in our study were small, possibility of exposure-outcome misclassification need to be considered as also indicated by selection bias within our tattooed population in regards to dose-response relationships. Such biases can only be properly addressed in prospective epidemiological cohort analyses, such as of the CRABAT cohort, that delivered the underlying data for this analysis, linking current tattoo exposure to future cancer risks. In any case, any protective effect of tattoos against skin cancer, if holding true, does not imply that tattoos are safe for other health outcomes as potential mechanisms of protection would be likely specific to skin cancer.

## Data availability

In accordance with the Constances Charter, de-identified participant data from the Constances cohort are available to researchers who meet the legal and ethical requirements set by the French National Commission governing data privacy laws. International researchers can access the dataset at https://www.constances.fr/en/scientific-area/access-to-constances-2/. Additionally, all study materials, including the study protocol and data dictionary of the Constances cohort, are freely accessible. All data is analysed on the secured CASD (Centre d’Access Securise aux Donnees) server. As this server does not allow export of data or codes, stored on the server, access to the codes can only be guaranteed after data access request. For further questions, please contact Milena Foerster.

## Author Contributions

Tingting Mo, PhD (Data curation; Formal analysis; Investigation; Methodology; Visualization; Validation, Writing—original draft; Writing—review & editing), Marie Zins, MD, PhD, Prof (Conceptualisation, Investigation, Project administration, Resources, Writing—review and editing), Marcel Goldberg, MD, PhD, Prof (Conceptualisation, Investigation, Project administration, Resources, Writing—review and editing), Céline Ribet, PhD (Investigation, Project administration, Resources), Sofiane Kab, PhD (Investigation, Project administration, Resources, Software), Ines Schreiver, PhD (Conceptualisation, Writing—original draft, Writing—review and editing), Katherina Siewert, PhD (Conceptualisation, Writing—original draft, Writing—review and editing), Khaled Ezzedine, MD, PhD, Prof (Conceptualisation, Investigation, Project administration, Resources, Writing—review and editing), Joachim Schuz, PhD (Conceptualisation, Funding acquisition, Investigation, Methodology, Writing—review and editing), Milena Foerster, PhD (Conceptualisation, Data curation, Funding acquisition, Investigation, Methodology, Project administration, Supervision, Writing—original draft; Writing— review & editing).

## Ethics approval

The CRABAT study received additional approval by Ethics Committee of the International Agency for research on Cancer (IEC 22-02), and was authorised by the French Data Protection Authority (“Commission Nationale de l’Informatique et des Libertés”, CNIL) (#22015584). The Constances study was approved by the Institutional Review Board (IRB) of the French Institute of Health (Inserm) (Opinion no. 01-011, then no. 21-842), and authorised by the CNIL (Authorization #910486).

## Disclaimer

Where authors are identified as personnel of the International Agency for Research on Cancer/World Health Organization, the authors alone are responsible for the views expressed in this article and they do not necessarily represent the decisions, policy, or views of International Agency for Research on Cancer /World Health Organization.

