## Supplementary materials for "Tattoos and risk of cutaneous melanoma and non-melanoma skin cancer in France"

Tingting Mo, PhD,1 Marie Zins, MD, PhD, Prof., 2 Marcel Goldberg, MD, PhD, Prof.,2 Céline Ribet, PhD,2 Sofiane Kab, PhD,2 Ines Schreiver, PhD,3 Katherina Siewert, PhD, Prof.3, Khaled Ezzedine, MD, PhD, Prof.,4,5 Joachim Schüz, PhD,1 Milena Foerster, PhD1*

1 Environment and Lifestyle Epidemiology Branch, International Agency for Research on Cancer, Lyon, France
2 Population-based Cohorts Unit, INSERM UMS 11, Paris Saclay University, Paris, France
3 Dermatotoxicology Study Centre, German Federal Institute for Risk Assessment (BfR), Berlin, Germany
4 Department of Dermatology, University Hospital Henri-Mondor, University of Paris Est-Créteil, Créteil, France
5 Epidemiology in Dermatology and Evaluation of Therapeutics, EA7379, Paris-Est University, Université Paris-Est Créteil (UPEC), Créteil, France

*Correspondence to: Milena Foerster, PhD, Environment and Lifestyle Epidemiology Branch, International Agency for Research on Cancer, 25 Avenue Tony Garnier, 69007 Lyon, France (

Abstract
Background: With the increasing popularity of decorative tattooing, which entails the intradermal injection of inks that may contain carcinogens, investigating the related potential skin cancer risk is a public health priority.
Methods: We used data from the Cancer Risk Attributable with the Body Art of Tattooing (CRABAT) study, nested in the French national cohort Constances (adults aged 18-69 years recruited in 2012–2018). Tattoo exposure was collected in 2020–23. Skin cancers overall, cutaneous melanoma (CM), and non-melanoma skin cancer (NMSC) diagnosed during 2007–21 were retrieved from national health insurance data. As exposure information was collected after possible disease ascertainment, risks of skin cancer with prior tattoo exposure were assessed using (i) Logistic regression and (ii) retrospective cohort analyses using Cox proportional hazards model.
Results: Among 111 074 participants, 1789 skin cancers (1.6%) were recorded (693 CM, 1096 NMSC). No association was found between binary tattoo exposure and any skin cancer type. In logistic regression, tattoo body surface >2 hand palms was associated with lower overall skin cancer risk (OR = 0.21, 95% CI: 0.05–0.83; reference no tattoos). This association was not significant in the Cox model, but the suggestive dose–response relationship remained, with HRs of 1.14, 0.60, and 0.26 for tattoo body surface of 0–1, 1–2, and >2 hand palms, respectively.
Conclusion: Large tattoo surfaces were tentatively associated with reduced overall skin cancer risk. While these finding merits further research, small case numbers and the retrospectively collected data might have biased the results.

**Supplementary Methods**

S1 Imputation for date of first tattoo

S2 Covariates selection

**Tables**

**Table S1.** Odds ratios (ORs) and confidence intervals (CIs) from logistic regression models of tattoo status associated with potential predictors for imputing age at first tattoo.

**Table S2.** Coefficients (β) and confidence intervals (CIs) from linear regression models of age at first tattoo associated with potential predictors for imputing age at first tattoo.

**Table S3.** Odds ratios (ORs) and confidence intervals (CIs) for overall skin cancer for participants with tattoo versus those without, with different levels of adjustment.

**Table S4.** Additional skin characteristics of the CRABAT participants not included for statistical analyses, for tattooed and non–tattooed individuals, and for tattooed individuals by tattooed body surface.

**Table S5.** Sociodemographic characteristics in 111074 eligible participants of the CRABAT study, by EpiTAT questionnaire respondent status.

**Table S6.** Skin characteristics in 111074 eligible participants of the CRABAT study, by EpiTAT questionnaire respondent status.

**Table S7.** Odds ratios (ORs) and confidence intervals (CIs) of multivariate logistic models with and without tattoo to explore independency of the tattoo exposure from other covariates.

**Table S8.** Hazard ratios (HRs) and confidence intervals (CIs) of Cox proportional hazard regression for the association of different tattoo exposure and risk of overall skin cancer, melanoma, and non-melanoma, in the whole population with study entry date corresponding to the individual Constances baseline date. Tattoo exposure was considered as time-varying exposure, meaning that an individual contributes person-years to the non-tattooed group until the date of first tattoo, when the individual changes exposure status and will be contributing to the exposed population thereafter. Date of first tattoo was imputed in ~40% of tattooed participants, i.e. those with only minimal exposure data (non-respondents to EpiTAT).

**Table S9.** Odds ratios (ORs) and confidence intervals (CIs) for the cross–sectional relationship of different tattoo exposure variables with overall skin cancer, melanoma, and non–melanoma in the population over 40 at first exposure assessment (2020/21) in the CRABAT study.

**Table S10.** Hazard ratios (HRs) and confidence intervals (CIs) of Cox proportional hazard regression for the association of different tattoo exposure and risk of overall skin cancer, melanoma, and non-melanoma, in the population over 40 at exposure assessment (2020/21) with study entry date on 1 Jan, 2007 (start of outcome assessment via French health database). Tattoo exposure was considered as time-varying exposure, meaning that an individual contributes person-years to the non-tattooed group until the date of first tattoo, when the individual changes exposure status and will be contributing to the exposed population thereafter. Date of first tattoo was imputed in ~40% of tattooed participants, i.e. those with only minimal exposure data (non-respondents to EpiTAT).

**Table S11.** Hazard ratios (HRs) and confidence intervals (CIs) of Cox proportional hazard regression for the association of different tattoo exposure and risk of overall skin cancer, melanoma, and non-melanoma among participants who provided first tattoo date with study entry date on 1 Jan, 2007 (start of outcome assessment via French health database). Tattoo exposure was considered as time-varying exposure, meaning that an individual contributes person-years to the non-tattooed group until the date of first tattoo, when the individual changes exposure status and will be contributing to the exposed population thereafter.

**Figures**

Figure S1. Directed acyclic graph (DAG) demonstrating covariate selection to estimate the association of tattoo exposure and skin cancer (including overall skin cancer, melanoma, and non-melanoma skin cancer). Tattoo is exposure and skin cancer is the outcome. Red circles are ancestor of exposure and outcome, while blue circles are ancestor of outcome.

**Supplementary Methods**

***S1 Imputation for date of first tattoo***

To account for the time-varying tattoo status, we split the dataset according to the first date of tattoo. In this study, 40% of the tattooed participants did not answer the detailed EpiTAT questionnaire thus did not provide date of first tattoo. As sensitivity analysis, we imputed these missing values using the following strategy. Potential predictors for being tattooed or not were selected via literature and extracted from the Constances baseline data. These included sex, age, number of sexual partners, homosexual intercourse amongst men, alcohol drinking status, smoking status, education, household disposable income, barriers to healthcare, and current occupation. We first tested their associations with tattoo status (yes, no) using logistic regression models (Table S1), then with age of first tattoo using linear regression models (Table S2). As a result, the variable barriers to healthcare was excluded as it was not associated with age of first tattoo (Table S1-S2). The factors left were used as predictors for imputation using the following commands in STATA, noted that the age of first tattoo was set to be at least 16 years-old and younger that the age at baseline when participants were first asked questions about tattoo:

mi impute chained (ologit) education income ///

(mlogit, augment) drinking smoking job ///

(truncreg, ll(16) ul(age_bl)) age_at_tattoo ///

=i.sex age i.nunber_sexual_partner i.sex_with_men skin if tattooed==1, add(10) rseed(123)

Using the imputed age of first tattoo, we generated the date of first tattoo which was used for stsplit (STATA).

**Table S1.** Odds ratios (ORs) and confidence intervals (CIs) from logistic regression models of tattoo status associated with potential predictors for imputing age at first tattoo.

| Variables | Univariate model[a] | | Multivariate model[a] | |
|---|---|---|---|---|
| | OR (95% CI) | P-value | OR (95% CI) | P-value |
| **Sex** | | | | |
| Female | ref. | | ref. | |
| Male | 0.69 (0.67-0.72) | <0.05 | 1.66 (1.57-1.75) | <0.05 |
| **Age at baseline** | 0.95 (0.95-0.95) | <0.05 | 3.08 (2.92-3.25) | <0.05 |
| **Number of sexual partners** | | | | |
| NA/never had sex | 0.76 (0.71-0.82) | <0.05 | 0.95 (0.86-1.04) | insignificant |
| 1 | ref. | | ref. | |
| 2-5 | 0.54 (0.51-0.58) | <0.05 | 0.64 (0.59-0.69) | <0.05 |
| 6-10 | 1.46 (1.38-1.54) | <0.05 | 1.42 (1.34-1.52) | <0.05 |
| >10 | 1.87 (1.77-1.99) | <0.05 | 1.98 (1.85-2.12) | <0.05 |
| Prefer not to answer | 1.01 (0.96-1.06) | insignificant | 1.30 (1.22-1.39) | <0.05 |
| **Men sleeping with men** | | | | |
| Women/heterosexual men | ref. | | ref. | |
| Yes | 1.32 (1.20-1.46) | <0.05 | 1.30 (1.16-1.47) | <0.05 |
| Unknown | 0.60 (0.49-0.75) | <0.05 | 0.76 (0.58-0.99) | <0.05 |
| **Alcohol drinking status** | | | | |
| Abstinent | 1.02 (0.97-1.07) | insignificant | 0.90 (0.85-0.95) | <0.05 |
| Abuse | 1.30 (1.22-1.38) | <0.05 | 1.14 (1.05-1.23) | <0.05 |
| Dependence | 1.29 (1.20-1.40) | <0.05 | 1.01 (0.92-1.10) | insignificant |
| Neither abuse nor dependence | ref. | | ref. | |
| **Smoking status** | | | | |
| Never | ref. | | ref. | |
| Former | 1.58 (1.51-1.65) | <0.05 | 1.80 (1.71-1.90) | <0.05 |
| Current | 3.18 (3.02-3.34) | <0.05 | 2.09 (1.97-2.22) | <0.05 |
| **Education[b]** | | | | |
| No diploma | 1.22 (1.16-1.28) | <0.05 | 2.21 (2.06-2.37) | <0.05 |
| High school degree | 1.43 (1.36-1.51) | <0.05 | 1.49 (1.39-1.58) | <0.05 |
| Bachelor degree | ref. | | ref. | |
| Master degree and higher | 0.57 (0.54-0.60) | <0.05 | 0.56 (0.52-0.60) | <0.05 |
| **Household disposable income** | | | | |
| ≤2100 € | 2.89 (2.73-3.06) | <0.05 | 1.14 (1.06-1.22) | <0.05 |
| 2100-2800€ | 2.16 (2.03-2.30) | <0.05 | 1.17 (1.09-1.26) | <0.05 |
| 2800-4200€ | 2.00 (1.89-2.10) | <0.05 | 1.24 (1.17-1.31) | <0.05 |
| ≥4200€ | ref. | | ref. | |
| **Barriers to healthcare[c]** | | | | |
| Yes | 1.69 (1.62-1.77) | <0.05 | 1.21 (1.14-1.27) | <0.05 |
| No | ref. | | ref. | |
| **Occupation** | | | | |
| Farmer/the spouse | 2.33 (1.79-3.03) | <0.05 | 1.20 (0.88-1.64) | insignificant |
| Craftsman, shopkeeper… | 2.19 (1.91-2.50) | <0.05 | 1.40 (1.19-1.63) | <0.05 |
| Higher intellectual profession | ref. | | ref. | |
| Intermediate profession | 1.66 (1.57-1.75) | <0.05 | 1.22 (1.14-1.31) | <0.05 |
| Employee | 3.08 (2.92-3.25) | <0.05 | 1.43 (1.33-1.55) | <0.05 |
| Manual worker | 3.31 (3.08-3.56) | <0.05 | 1.79 (1.62-1.99) | <0.05 |
| Has never worked | 3.13 (2.73-3.59) | <0.05 | 0.56 (0.46-0.68) | <0.05 |
| Other | 2.20 (1.63-2.98) | <0.05 | 1.13 (0.81-1.59) | insignificant |

[a] Univariate models explored the association of a single variables with the outcome, while multivariate models adjusted for all listed variables.
[b] Original categorisation according to French education system, i.e. "high school degree" equals the French "baccalauréat général" after 12 years of schooling, "bachelor degree" equals a university degree obtained after up to four years ("bac +2/3/4"), and "master degree" equals a university degree obtained after at least five years ("bac +5 et superieur").
[c] Dichotomous variable composed from three variables enquiring whether in the last 12 months the participant or a family member had to forego healthcare due to (1) financial problems, (2) long distance to medical services or (3) too long waiting times for an appointment· Variable was coded "yes" if any of these were answered with "yes" in at least one of (up to) six follow–up questionnaires.

**Table S2**. Coefficients (β) and confidence intervals (CIs) from linear regression models of age at first tattoo associated with potential predictors for imputing age at first tattoo.

| Variables | Univariate model[a] | | Multivariate model[a] | |
|---|---|---|---|---|
| | β (95% CI) | P-value | β (95% CI) | P-value |
| **Sex** | | | | |
| Female | ref. | | ref. | |
| Male | 4.09 (3.56, 4.62) | <0.05 | -0.20 (-0.93, 0.53) | insignificant |
| **Age at baseline** | 0.84 (0.82, 0.85) | <0.05 | -1.32 (-2.04, -0.61) | <0.05 |
| **Number of sexual partners** | | | | |
| NA/never had sex | 3.69 (2.62, 4.76) | <0.05 | -0.31 (-1.01, 0.39) | insignificant |
| 1 | ref. | | ref. | |
| 2-5 | -0.08 (-1.05, 0.89) | insignificant | 0.68 (0.09, 1.27) | <0.05 |
| 6-10 | 0.13 (-0.62, 0.87) | insignificant | -0.50 (-0.95, -0.06) | <0.05 |
| >10 | -0.28 (-1.07, 0.52) | insignificant | -1.31 (-1.80, -0.82) | <0.05 |
| Prefer not to answer | 4.37 (3.61, 5.14) | <0.05 | 0.05 (-0.44, 0.53) | insignificant |
| **Men sleeping with men** | | | | |
| Women/heterosexual men | ref. | | ref. | |
| Yes | 1.46 (0.12, 2.81) | <0.05 | 0.60 (-0.23, 1.42) | insignificant |
| Unknown | 4.96 (1.50, 8.42) | <0.05 | -1.61 (-3.91, 0.69) | insignificant |
| **Alcohol drinking status** | | | | |
| Abstinent | -1.86 (-2.56, -1.16) | <0.05 | -0.34 (-0.76, 0.09) | insignificant |
| Abuse | -0.26 (-1.20, 0.68) | insignificant | -0.43 (-1.00, 0.13) | insignificant |
| Dependence | -0.87 (-1.98, 0.23) | insignificant | 0.25 (-0.42, 0.91) | insignificant |
| Neither abuse nor dependence | ref. | | ref. | |
| **Smoking status** | | | | |
| Never | ref. | | ref. | |
| Former | 3.80 (3.21, 4.38) | <0.05 | -0.38 (-0.75, 0.00) | <0.05 |
| Current | -0.59 (-1.27, 0.08) | insignificant | -0.70 (-1.12, -0.28) | <0.05 |
| **Education[b]** | | | | |
| No diploma | 7.86 (7.20, 8.53) | <0.05 | 0.13 (-0.37, 0.64) | insignificant |
| High school degree | 0.85 (0.17, 1.54) | <0.05 | 0.33 (-0.13, 0.79) | insignificant |
| Bachelor degree | ref. | | ref. | |
| Master degree and higher | -1.08 (-1.78, -0.38) | <0.05 | 0.45 (-0.04, 0.93) | insignificant |
| **Household disposable income** | | | | |
| ≤2100 € | -1.66 (-2.45, -0.88) | <0.05 | 1.12 (0.60, 1.64) | <0.05 |
| 2100-2800€ | -0.80 (-1.65, 0.06) | insignificant | 0.88 (0.35, 1.41) | <0.05 |
| 2800-4200€ | -1.47 (-2.18, -0.75) | <0.05 | 0.28 (-0.16, 0.72) | insignificant |
| ≥4200€ | ref. | | ref. | |
| **Barriers to healthcare[c]** | | | | |
| Yes | 0.15 (-0.46, 0.77) | insignificant | NA | |
| No | ref. | | NA | |
| **Occupation** | | | | |
| Farmer/the spouse | 1.29 (-2.46, 5.04) | insignificant | 0.26 (-2.04, 2.56) | insignificant |
| Craftsman, shopkeeper… | 0.25 (-1.63, 2.14) | insignificant | -1.14 (-2.30, 0.02) | insignificant |
| Higher intellectual profession | ref. | | | |
| Intermediate profession | -0.20 (-0.93, 0.53) | insignificant | -0.67 (-1.17, -0.18) | <0.05 |
| Employee | -1.32 (-2.04, -0.61) | <0.05 | -0.59 (-1.15, -0.04) | <0.05 |
| Manual worker | 2.35 (1.33, 3.37) | <0.05 | -1.78 (-2.53, -1.02) | <0.05 |
| Has never worked | -12.05 (-13.91, -10.19) | <0.05 | 3.34 (1.97, 4.72) | <0.05 |
| Other | 0.03 (-4.22, 4.29) | insignificant | 1.09 (-1.42, 3.60) | insignificant |

[a] Univariate models explored the association of a single variables with the outcome, while multivariate models adjusted for all listed variables.
[b] Original categorisation according to French education system, i.e. "high school degree" equals the French "baccalauréat général" after 12 years of schooling, "bachelor degree" equals a university degree obtained after up to four years ("bac +2/3/4"), and "master degree" equals a university degree obtained after at least five years ("bac +5 et superieur").
[c] Dichotomous variable composed from three variables enquiring whether in the last 12 months the participant or a family member had to forego healthcare due to (1) financial problems, (2) long distance to medical services or (3) too long waiting times for an appointment· Variable was coded "yes" if any of these were answered with "yes" in at least one of (up to) six follow–up questionnaires. Barriers to healthcare was not included in the multivariate model as it was not significant in the univariate model.

***S2 Covariates selection***

Factors that potentially associated with both tattoo and skin cancer were first selected based on previous literature, including age at baseline, sex, education (no diploma, high school degree, bachelor degree, master degree and higher), household disposable income (≤2100€, 2100 to <2800€, 2800 to <4200€, ≥4200€), smoking status (never smoked, former smoker, current smoker), body mass index (BMI), skin type (vary fair [white], very faire and may be tanned, moderately fair, matt, dark brown), number of moles and freckles (none, very few, few, many), maximum intensity of tan (absent, light, bright, dark, very dark), frequency of sunburn during adolescence and during adulthood (never, 1–5 times, 6–10 times, every summer, I don't know), barriers to healthcare (not impaired, impaired), physical activity outside of work (never or nearly never, rarely [<2h/day], frequently [2–4h/day], always or nearly always), and occupation (farmer/the spouse, craftsman or shopkeeper, higher intellectual pression, intermediate pr0fession, employee, manual worker, has never worked, other). These covariates were further tested by assessing how the factors changed the estimates when each was added to the logistic model in the aforementioned order starting with age at baseline and sex. Factors which did not change the point estimates and 95% confidence intervals ≥0.01 were excluded from the final models (Table S3).

**Table S3.** Odds ratios (ORs) and confidence intervals (CIs) for overall skin cancer for participants with tattoo versus those without, with different levels of adjustment.

| Model | OR (95% CI) |
|---|---|
| Age and sex | 0.80 (0.64-0.99) |
| + education level | 0.83 (0.67-1.04) |
| + household disposable income | 0.85 (0.68-1.06) |
| + smoking status | 0.86 (0.69-1.08) |
| + BMI | 0.87 (0.70-1.09) |
| + skin type | 0.91 (0.73-1.14) |
| + No. of moles | 0.91 (0.73-1.14) |
| + No. of freckle | 0.91 (0.73-1.14) |
| + maximum intensity of tan | 0.92 (0.73-1.15) |
| + frequency of sunburn during adolescence | 0.92 (0.74-1.15) |
| + frequency of sunburn during adulthood | 0.92 (0.74-1.15) |
| + barriers to healthcare | 0.92 (0.74-1.16) |
| + physical activity outside of work | 0.92 (0.74-1.16) |
| + occupation | 0.92 (0.74-1.16) |

**Table S4.** Additional skin characteristics of the CRABAT participants not included for statistical analyses, for tattooed and non–tattooed individuals, and for tattooed individuals by tattooed body surface.

| Characteristic | Tattoo status, 2020 Constances respondents | | Tattoo size, among all tattooed[a] | | | |
|---|---|---|---|---|---|---|
| | Tattooed | Not tattooed | 0–1 hand surface | 1–2 hand surface | >2 hand surfaces | Tattooed, size missing |
| | N (% among full sample, in category) | | N (% among the tattooed categories) | | | |
| No of participants (% row) | 12669 (11.5%) | 98405 (89.5%) | 6712 (53.3%) | 2001 (15.9%) | 1633 (13.0%) | 2323 (18.5%) |
| No of moles, N (%) known [b] | 7315 (57.7%) | 63431 (64.5%) | 3987 (59.4%) | 1224 (61.2%) | 983 (60.2%) | 1121 (48.3%) |
| None | 524 (7.2%) | 7228 (11.4%) | 330 (8.3%) | 74 (6.0%) | 48 (4.9%) | 72 (6.4%) |
| Very few | 3439 (47.0%) | 33605 (53.0%) | 1917 (48.1%) | 548 (44.8%) | 422 (42.9%) | 552 (49.2%) |
| Few | 2452 (33.5%) | 17114 (27.0%) | 1285 (32.2%) | 429 (35.0%) | 369 (37.5%) | 369 (32.9%) |
| Many | 900 (12.3%) | 5484 (8.6%) | 455 (11.4%) | 173 (14.1%) | 144 (14.6%) | 128 (11.4%) |
| No. of freckles, N (%) known [b] | 7377 (58.2%) | 64128 (65.2%) | 4023 (59.9%) | 1240 (62.0%) | 985 (60.3%) | 1129 (48.6%) |
| None | 4166 (56.5%) | 38206 (59.6%) | 2218 (55.1%) | 727 (58.6%) | 575 (58.4%) | 646 (57.2%) |
| Very few | 2162 (29.3%) | 17490 (27.3%) | 1225 (30.4%) | 335 (27.0%) | 268 (27.2%) | 334 (29.6%) |
| Few | 801 (10.9%) | 6484 (10.1%) | 443 (11.0%) | 145 (11.7%) | 102 (10.4%) | 111 (9.8%) |
| Many | 248 (3.4%) | 1948 (3.0%) | 137 (3.4%) | 33 (2.7%) | 40 (4.1%) | 38 (3.4%) |
| Sunburn during adulthood, N (%) known[b] | 6619 (52.2%) | 56063 (57.0%) | 3590 (53.5%) | 1127 (56.3%) | 898 (55.0%) | 1004 (43.2%) |
| Never | 173 (2.6%) | 2292 (4.1%) | 92 (2.6%) | 29 (2.6%) | 29 (3.2%) | 23 (2.3%) |
| 1–5 times | 2948 (44.5%) | 24571 (43.8%) | 1593 (44.4%) | 490 (43.5%) | 409 (45.5%) | 456 (45.4%) |
| 6–10 times | 1492 (22.5%) | 13779 (24.6%) | 859 (23.9%) | 254 (22.5%) | 184 (20.5%) | 195 (19.4%) |
| Every summer | 1509 (22.8%) | 10387 (18.5%) | 781 (21.8%) | 279 (24.8%) | 203 (22.6%) | 246 (24.5%) |
| I don't know | 497 (7.5%) | 5034 (9.0%) | 265 (7.4%) | 75 (6.7%) | 73 (8.1%) | 84 (8.4%) |

[a] Tattoo surface retrieved from second phase exposure assessment in 2023 (EpiTAT exposure questionnaire). In addition, participants who only took part in the first phase exposure assessment in 2020 (Constances follow-up questionnaire) and who therein reported tattoo surface of "≤ 1 hand surface" (vs ">1 hand surface") were added to the lowest exposure category.
[b] N (%) known indicates the number and percentage of non-missing values for each variable.

**Table S5.** Sociodemographic characteristics in 111074 eligible participants of the CRABAT study, by EpiTAT questionnaire respondent status.

| Characteristics | Not tattooed | Tattooed | | p-value[a] |
|---|---|---|---|---|
| | | EpiTAT responders | EpiTAT non-responders | |
| No. (%) of participants | 98405 (88.6%) | 7553 (6.8%) | 5116 (4.6%) | |
| Sex | | | | 0.008 |
| Male | 45606 (46.3%) | 2753 (36.4%) | 1983 (38.8%) | |
| Female | 52799 (53.7%) | 4800 (63.6%) | 3133 (61.2%) | |
| Age at baseline, median (IQR) | 51 (40, 61) | 40 (32, 48) | 40 (32, 49) | 0.60 |
| BMI, kg/m$^2$, N (%) known [b] | 97122 (98.7%) | 7464 (98.8%) | 5067 (99.0%) | |
| Median (IQR) | 24.2 (21.8, 27.0) | 23.9 (21.5, 27.0) | 24.2 (21.8, 27.5) | <0.001 |
| Smoking status, N (%) known [b] | 97061 (98.6%) | 7489 (99.2%) | 5028 (98.3%) | <0.001 |
| Never | 49340 (50.8%) | 2741 (36.6%) | 1555 (30.9%) | |
| Former | 35753 (36.8%) | 2963 (39.6%) | 1948 (38.7%) | |
| Current | 11968 (12.3%) | 1785 (23.8%) | 1525 (30.3%) | |
| Education, N (%) known[b,c] | 96596 (98.2%) | 7461 (98.8%) | 4998 (97.7%) | <0.001 |
| No diploma/bac | 19403 (20.1%) | 1624 (21.8%) | 1480 (29.6%) | |
| Bac or equal | 14047 (14.5%) | 1504 (20.2%) | 1138 (22.8%) | |
| Bac+2/3/4 | 35272 (36.5%) | 2946 (39.5%) | 1684 (33.7%) | |
| Bac+5 and more | 27874 (28.9%) | 1387 (18.6%) | 696 (13.9%) | |
| Household disposable income, N (%) known [b] | 92188 (93.7%) | 7093 (93.9%) | 4704 (91.9%) | <0.001 |
| ≤2100 € | 15104 (16.4%) | 1692 (23.9%) | 1404 (29.8%) | |
| 2100-2800€ | 13370 (14.5%) | 1221 (17.2%) | 824 (17.5%) | |
| 2800-4200€ | 30374 (32.9%) | 2638 (37.2%) | 1656 (35.2%) | <0.001 |
| ≥4200€ | 33340 (36.2%) | 1542 (21.7%) | 820 (17.4%) | |
| Barriers to healthcare[d] | | | | 0.69 |
| Yes | 83316 (84.7%) | 5793 (76.7%) | 3908 (76.4%) | |
| No | 15089 (15.3%) | 1760 (23.3%) | 1208 (23.6%) | |

[a] Characteristics were compared between EpiTAT responders versus non-responders

[b] N (%) known indicates the number and percentage of non-missing values for each variable.

[c] Original categorisation according to French education system, i.e. "high school degree" equals the French "baccalauréat général" after 12 years of schooling, "bachelor degree" equals a university degree obtained after up to four years ("bac +2/3/4"), and "master degree" equals a university degree obtained after at least five years ("bac +5 et superieur").

[d] Dichotomous variable composed from three variables enquiring whether in the last 12 months the participant or a family member had to forego healthcare due to (1) financial problems, (2) long distance to medical services or (3) too long waiting times for an appointment· Variable was coded "yes" if any of these were answered with "yes" in at least one of (up to) six follow–up questionnaires.

**Table S6.** Skin characteristics in 111074 eligible participants of the CRABAT study, by EpiTAT questionnaire respondent status.

| Characteristics | Not tattooed | Tattooed | | |
|---|---|---|---|---|
| | | EpiTAT responders | EpiTAT non-responder | p-value[a] |
| No. of participants | 98405 | 7553 | 5116 | |
| Maximum intensity of tan, N (%) known [b] | 63627 (64.7%) | 4726 (62.6%) | 2601 (50.8%) | 0.16 |
| Absent | 988 (1.6%) | 45 (1.0%) | 27 (1.0%) | |
| Light | 12062 (19.0%) | 646 (13.7%) | 375 (14.4%) | |
| Bright | 19685 (30.9%) | 1343 (28.4%) | 667 (25.6%) | |
| Dark | 28995 (45.6%) | 2455 (51.9%) | 1397 (53.7%) | |
| Very dark | 1897 (3.0%) | 237 (5.0%) | 135 (5.2%) | |
| No. of moles, N (%) known [b] | 63431 (64.5%) | 4726 (62.6%) | 2589 (50.6%) | <0.001 |
| None | 7228 (11.4%) | 308 (6.5%) | 216 (8.3%) | |
| Very few | 33605 (53.0%) | 2174 (46.0%) | 1265 (48.9%) | |
| Few | 17114 (27.0%) | 1628 (34.4%) | 824 (31.8%) | |
| Many | 5484 (8.6%) | 616 (13.0%) | 284 (11.0%) | |
| No. of freckles, N (%) known [b] | 64128 (65.2%) | 4761 (63.0%) | 2616 (51.1%) | 0.43 |
| None | 38206 (59.6%) | 2666 (56.0%) | 1500 (57.3%) | |
| Very few | 17490 (27.3%) | 1397 (29.3%) | 765 (29.2%) | |
| Few | 6484 (10.1%) | 537 (11.3%) | 264 (10.1%) | |
| Many | 1948 (3.0%) | 161 (3.4%) | 87 (3.3%) | |
| Skin type, N (%) known[b,c] | 62944 (64.0%) | 4701 (62.2%) | 2565 (50.1%) | 0.012 |
| Very fair, white | 1660 (2.6%) | 124 (2.6%) | 88 (3.4%) | |
| Very fair, and may be tanned | 9479 (15.1%) | 823 (17.5%) | 407 (15.9%) | |
| Moderately fair | 34339 (54.6%) | 2488 (52.9%) | 1349 (52.6%) | |
| Matt | 16500 (26.2%) | 1187 (25.2%) | 655 (25.5%) | |
| Dark brown | 966 (1.5%) | 79 (1.7%) | 66 (2.6%) | |
| Times of sunburn during adolescence, N (%) known [b] | 63303 (64.3%) | 4718 (62.5%) | 2595 (50.7%) | 0.016 |
| Never | 6783 (10.7%) | 456 (9.7%) | 302 (11.6%) | |
| 1-5 times | 17572 (27.8%) | 1276 (27.0%) | 712 (27.4%) | |
| 6-10 times | 13797 (21.8%) | 1022 (21.7%) | 526 (20.3%) | |
| Every summer | 18440 (29.1%) | 1501 (31.8%) | 772 (29.7%) | |
| I don't know | 6711 (10.6%) | 463 (9.8%) | 283 (10.9%) | |
| Times of sunburn during adulthood, N (%) known [b] | 56063 (57.0%) | 4310 (57.1%) | 2309 (45.1%) | 0.55 |
| Never | 2292 (4.1%) | 106 (2.5%) | 67 (2.9%) | |
| 1-5 times | 24571 (43.8%) | 1914 (44.4%) | 1034 (44.8%) | |
| 6-10 times | 13779 (24.6%) | 992 (23.0%) | 500 (21.7%) | |
| Every summer | 10387 (18.5%) | 983 (22.8%) | 526 (22.8%) | |
| I don't know | 5034 (9.0%) | 315 (7.3%) | 182 (7.9%) | |
| Skin cancer incidence | | | | |
| Overall skin cancer | 1702 (1.7%) | 38 (0.5%) | 49 (1.0%) | |
| Melanoma | 659 (0.7%) | 16 (0.2%) | 18 (0.4%) | |
| Non-melanoma | 1043 (1.1%) | 22 (0.3%) | 31 (0.6%) | |

[a] Characteristics were compared between EpiTAT responders versus non-responders
[b] N (%) known indicates the number and percentage of available data for each variable with missing values.
[c] Categorization according to the Fitzpatrick scale. Individuals with Fitzpatrick type IV (black skin, n=292) were excluded from the study because of their very low skin cancer risk.

**Table S7**. Odds ratios (ORs) and confidence intervals (CIs) of multivariate logistic models with and without including tattooed body surface to explore independency of the tattoo exposure from other covariates.

| Variables | OR (95% CI)[a] | |
|---|---|---|
| | **Without tattooed body surface** | **With tattooed body surface** |
| Age at Constances baseline | 1.08 (1.08, 1.09) | 1.08 (1.08, 1.09) |
| Sex | | |
| Male | 1.20 (1.08, 1.33) | 1.20 (1.08, 1.33) |
| Female | ref | ref |
| Smoking status | | |
| Never | ref | ref |
| Former | 1.00 (0.91, 1.11) | 1.01 (0.91, 1.12) |
| Current | 0.84 (0.70, 1.00) | 0.83 (0.69, 1.00) |
| BMI | | |
| <18 | 1.37 (0.90, 2.07) | 1.38 (0.91, 2.09) |
| 18 to <20 | 1.13 (0.93, 1.38) | 1.12 (0.92, 1.37) |
| 20 to <25 | ref | ref |
| 25 to <30 | 0.92 (0.82, 1.02) | 0.92 (0.82, 1.03) |
| 30+ | 0.79 (0.67, 0.93) | 0.79 (0.67, 0.93) |
| Barriers to healthcare[b] | | |
| Yes | ref | ref |
| No | 0.95 (0.82, 1.09) | 0.94 (0.82, 1.09) |
| Household disposable income | | |
| ≤2100 € | 0.75 (0.63, 0.89) | 0.75 (0.63, 0.89) |
| 2100-2800€ | 0.88 (0.75, 1.03) | 0.88 (0.75, 1.03) |
| 2800-4200€ | 0.82 (0.72, 0.92) | 0.81 (0.72, 0.92) |
| ≥4200€ | ref | ref |
| Education[c] | | |
| No diploma/bac | 0.87 (0.77, 1.00) | 0.89 (0.78, 1.01) |
| Bac or equal | 0.96 (0.83, 1.11) | 0.96 (0.83, 1.11) |
| Bac+2/3/4 | ref | ref |
| Bac+5 and more | 0.89 (0.78, 1.01) | 0.89 (0.78, 1.01) |
| Skin type[d] | | |
| Very fair, white | 1.52 (1.15, 2.02) | 1.50 (1.13, 2.00) |
| Very fair, and may be tanned | 1.59 (1.37, 1.85) | 1.59 (1.37, 1.85) |
| Moderately fair | 0.66 (0.56, 0.78) | 0.66 (0.56, 0.78) |
| Matt | ref | ref |
| Dark brown | 0.59 (0.31, 1.12) | 0.59 (0.31, 1.13) |
| Maximum intensity of tan | | |
| Absent | 2.21 (1.61, 3.03) | 2.22 (1.62, 3.05) |
| Light | 1.24 (1.06, 1.45) | 1.24 (1.06, 1.45) |
| Bright | 1.09 (0.94, 1.26) | 1.08 (0.93, 1.25) |
| Dark | ref | ref |
| Very dark | 0.84 (0.51, 1.38) | 0.86 (0.52, 1.41) |
| Times of sunburn during adolescence | | |
| Never | 0.49 (0.39, 0.62) | 0.49 (0.39, 0.62) |
| 1-5 times | 0.68 (0.59, 0.79) | 0.68 (0.59, 0.79) |
| 6-10 times | 0.73 (0.63, 0.85) | 0.73 (0.63, 0.85) |
| Every summer | ref | ref |
| I don't know | 0.69 (0.57, 0.84) | 0.69 (0.57, 0.84) |

[a] Multivariate models included biological sex, age at Constance's baseline, education level, smoking status, BMI, household disposable income, skin type, maximum intensity of tan, frequency of sunburn during adolescence, and barriers to healthcare. Tattooed body surface was categorized as not tattooed, or 0-1, 1-2, >2 hand palms.
[b] Dichotomous variable composed from three variables enquiring whether in the last 12 months the participant or a family member had to forego healthcare due to (1) financial problems, (2) long distance to medical services or (3) too long waiting times for an appointment· Variable was coded "yes" if any of these were answered with "yes" in at least one of (up to) six follow–up questionnaires.
[c] Original categorisation according to French education system, i.e. "high school degree" equals the French "baccalauréat général" after 12 years of schooling, "bachelor degree" equals a university degree obtained after up to four years ("bac +2/3/4"), and "master degree" equals a university degree obtained after at least five years ("bac +5 et superieur").
[d] Categorization according to the Fitzpatrick scale. Individuals with Fitzpatrick type IV (black skin, n=292) were excluded from the study because of their very low skin cancer risk.

**Table S8.** Hazard ratios (HRs) and confidence intervals (CIs) of Cox proportional hazard regression for the association of different tattoo exposure and risk of overall skin cancer, melanoma, and non-melanoma, in the whole population with study entry date corresponding to the individual Constances baseline date. Tattoo exposure was considered as time-varying exposure, meaning that an individual contributes person-years to the non-tattooed group until the date of first tattoo, when the individual changes exposure status and will be contributing to the exposed population thereafter. Date of first tattoo was imputed in ~40% of tattooed participants, i.e. those with only minimal exposure data (non-respondents to EpiTAT).

| | | Overall skin | | Melanoma | | Non-melanoma | |
|---|---|---|---|---|---|---|---|
| **Exposure** | **n/category[a]** | **Cases (% of category)** | **HR (95% CI)[b]** | **Cases (% of category)** | **HR (95% CI)[b]** | **Cases (% of category)** | **HR (95% CI)[b]** |
| **Tattooed** | | | | | | | |
| **N Total** | 110220 | 1008 (0.9%) | | 420 (0.4%) | | 588 (0.5%) | |
| No | 97602 | 961 (1.0%) | ref | 399 (0.4%) | ref | 562 (0.6%) | ref |
| Yes | 12618 | 47 (0.4%) | 0.99 (0.73-1.34) | 21 (0.2%) | 0.67 (0.39-1.15) | 26 (0.2%) | 1.00 (0.67-1.50) |
| **Tattooed body surface** | | | | | | | |
| **N Total** | 107905 | 996 (0.9%) | | 415 (0.4%) | | 581 (0.5%) | |
| Not tattooed | 97602 | 961 (1.0%) | ref | 399 (0.4%) | ref | 562 (0.6%) | ref |
| 0-1 hand palm | 6673 | 28 (0.4%) | 0.94 (0.64-1.38) | 15 (0.2%) | 1.13 (0.67-1.91) | 13 (0.2%) | 0.79 (0.45-1.38) |
| 1-2 hand palms | 1997 | 5 (0.3%) | 0.80 (0.33-1.94) | 1 (0.1%) | 0.35 (0.05-2.52) | 4 (0.2%) | 1.19 (0.44-3.20) |
| >2 hand palms | 1633 | 2 (0.1%) | 0.44 (0.11-1.76) | No obs | No obs | 2 (0.1%) | 0.82 (0.20-3.29) |
| ***Only amongst the tattooed :*** | | | | | | | |
| **Incremental among tattooed** | 10303 | 35 (0.3%) | 0.95 (0.84-1.07) | 16 (0.2%) | 0.67 (0.39-1.15) | 19 (0.2%) | 0.99 (0.91-1.08) |
| **Sun exposure of tattoo** | | | | | | | |
| **N Total** | 5961 | 15 (0.3%) | | 8 (0.1%) | | 7 (0.1%) | |
| Not sun exposed | 1025 | 2 (0.2%) | 0.32 (0.04-2.48) | 1 (0.1%) | 1.42 (0.11-19.13) | 1 (0.1%) | NA[c] |
| Sun exposed | 4936 | 13 (0.3%) | ref | 7 (0.1%) | ref | 6 (0.1%) | ref |

[a] Due to the different numbers of missing values for each tattoo–related variable, the number of participants (i.e. N Total for each tattoo exposure) were slightly different.

[b] Considering tattooed or not as time-varying, the dataset was split on date of first tattoo. Cox regression models were performed based on an age-time scale, with stratification of age at first exposure assessment in 2020/21 (<40, ≥40) and adjustment for biological sex, education level, smoking status, BMI, household disposable income, barriers to health care, maximum intensity of tan, skin type, frequency of sunburns during adolescence.

[c] NA=not available. The analysis could not be performed due to the limited sample size.

**Table S9.** Odds ratios (ORs) and confidence intervals (CIs) for the cross–sectional relationship of different tattoo exposure variables with overall skin cancer, melanoma, and non–melanoma in the population over 40 at first exposure assessment (2020/21) in the CRABAT study.

| Exposure variable | n/category[a] | Overall skin cancer | | Cutaneous melanoma | | Non–melanoma skin cancer | |
|---|---|---|---|---|---|---|---|
| | | Cases (% of category) | OR (95% CI)[b] | Cases (% of category) | OR (95% CI)[b] | Cases (% of category) | OR (95% CI)[b] |
| **Tattooed** | | | | | | | |
| **N Total** | 89844 | 1751 (1.9%) | | 673 (0.7%) | | 1078 (1.2%) | |
| No | 81760 | 1670 (2.1%) | ref | 643 (0.8%) | ref | 1027 (1.3%) | ref |
| Yes | 8084 | 81 (1.0%) | 0.93 (0.74-1.18) | 30 (0.4%) | 0.78 (0.54-1.14) | 51 (0.6%) | 1.05 (0.78-1.40) |
| **Tattooed body surface** | | | | | | | |
| **N Total** | 88584 | 1736 (2.0%) | | 670 (0.8%) | | 1078 (1.2%) | |
| Not tattooed | 81760 | 1670 (2.1%) | ref | 643 (0.8%) | ref | 1027 (1.3%) | ref |
| 0-1 hand palm | 4758 | 59 (1.3%) | 1.06 (0.81-1.39) | 25 (0.5%) | 1.04 (0.69-1.56) | 34 (0.7%) | 1.08 (0.76-1.53) |
| 1-2 hand palms | 1161 | 5 (0.4%) | 0.44 (0.18-1.06) | 2 (0.2%) | 0.38 (0.10-1.55) | 3 (0.3%) | 0.48 (0.15-1.50) |
| >2 hand palms | 905 | 2 (0.2%) | 0.24 (0.06-0.96) | No obs | No obs | 2 (0.2%) | 0.45 (0.11-1.80) |
| ***Only amongst the tattooed :*** | | | | | | | |
| Incremental among tattooed | 6824 | 66 (1.0%) | 0.85 (0.70-1.03) | 27 (0.4%) | 0.57 (0.30-1.07) | 39 (0.6%) | 0.92 (0.78-1.09) |
| **Sun exposure of tattoo** | | | | | | | |
| **N Total** | 3721 | 24 (0.6%) | | 11 (0.3%) | | 13 (0.3%) | |
| Not sun exposed | 800 | 8 (1.0%) | 1.54 (0.59-4.06) | 3 (0.4%) | 2.08 (0.43-10.08) | 5 (0.6%) | 0.95 (0.22-4.16) |
| Sun exposed | 2921 | 16 (0.5%) | ref | 8 (0.3%) | ref | 8 (0.3%) | ref |

[a] Due to the different numbers of missing values for each tattoo–related variable, the number of participants were slightly different.

[b] Model adjusted for biological sex, age at Constance's baseline, education level, smoking status, BMI, household disposable income, skin type, maximum intensity of tan, frequency of sunburn during adolescence, and barriers to healthcare.

**Table S10.** Hazard ratios (HRs) and confidence intervals (CIs) of Cox proportional hazard regression for the association of different tattoo exposure and risk of overall skin cancer, melanoma, and non-melanoma, in the population over 40 at first exposure assessment (2020/21) with study entry date on 1 Jan, 2007 (start of outcome assessment via French health database). Tattoo exposure was considered as time-varying exposure, meaning that an individual contributes person-years to the non-tattooed group until the date of first tattoo, when the individual changes exposure status and will be contributing to the exposed population thereafter. Date of first tattoo was imputed in ~40% of tattooed participants, i.e. those with only minimal exposure data (non-respondents to EpiTAT).

| | | **Overall skin cancer** | | **Cutaneous melanoma** | | **Non–melanoma skin cancer** | |
|---|---|---|---|---|---|---|---|
| **Exposure** | **n/category[a]** | **Cases (% of category)** | **HR (95% CI)[b]** | **Cases (% of category)** | **HR (95% CI)[b]** | **Cases (% of category)** | **HR (95% CI)[b]** |
| **Tattooed** | | | | | | | |
| **N Total** | 89844 | 1751 (1.9%) | | 673 (0.7%) | | 1078 (1.2%) | |
| No | 81760 | 1670 (2.0%) | ref | 643 (0.8%) | ref | 1027 (1.3%) | ref |
| Yes | 8084 | 81 (1.0%) | 1.04 (0.82-1.32) | 30 (0.4%) | 0.93 (0.63-1.37) | 51 (0.6%) | 1.11 (0.82-1.51) |
| **Tattooed body surface** | | | | | | | |
| **N Total** | 88584 | 1736 (1.9%) | | 670 (0.8%) | | 1066 (1.2%) | |
| Not tattooed | 81760 | 1670 (2.0%) | ref | 643 (0.8%) | ref | 1027 (1.3%) | ref |
| <1 hand palm | 4758 | 59 (1.2%) | 1.16 (0.88-1.53) | 25 (0.5%) | 1.21 (0.80-1.84) | 34 (0.7%) | 1.12 (0.78-1.62) |
| 1-2 hand palms | 1161 | 5 (0.4%) | 0.55 (0.23-1.33) | 2 (0.2%) | 0.52 (0.13-2.07) | 3 (0.3%) | No obs |
| >2 hand palms | 905 | 2 (0.2%) | 0.30 (0.07-1.19) | 0 (0.0%) | No obs | 2 (0.2%) | 0.53 (0.13-2.13) |
| ***Only amongst the tattooed :*** | | | | | | | |
| **Incremental among tattooed** | 6824 | 66 (1.0%) | 0.92 (0.80-1.04) | 27 (0.4%) | 0.78 (0.55-1.10) | 39 (0.6%) | 0.96 (0.85-1.08) |
| **Sun exposure of tattoo** | | | | | | | |
| **N Total** | 3721 | 24 (0.6%) | | 11 (0.3%) | | 13 (0.3%) | |
| Not sun exposed | 800 | 8 (1.0%) | 1.68 (0.64-4.41) | 3 (0.4%) | 2.26 (0.49-10.50) | 5 (0.6%) | 1.19 (0.30-4.71) |
| Sun exposed | 2921 | 16 (0.5%) | ref | 8 (0.3%) | ref | 8 (0.3%) | ref |

[a] Due to the different numbers of missing values for each tattoo–related variable, the number of participants (i.e. N Total for each tattoo exposure) were slightly different.
[b] Considering tattooed or not as time-varying, the dataset was split on date of first tattoo. Cox regression models were performed based on an age-time scale, with stratification of age at first exposure assessment in 2020/21 (<40, ≥40) and adjustment for biological sex, education level, smoking status, BMI, household disposable income, barriers to health care, maximum intensity of tan, skin type, frequency of sunburns during adolescence.

**Table S11.** Hazard ratios (HRs) and confidence intervals (CIs) of Cox proportional hazard regression for the association of different tattoo exposure and risk of overall skin cancer, melanoma, and non-melanoma among participants who provided first tattoo date with study entry date on 1 Jan, 2007 (start of outcome assessment via French health database). Analyses restricted to the participants with known first tattoo date, i.e. those who responded to the EpiTAT questionnaire in 2023.Tattoo exposure was considered as time-varying exposure, meaning that an individual contributes person-years to the non-tattooed group until the date of first tattoo, when the individual changes exposure status and will be contributing to the exposed population thereafter.

| | | Overall skin | | Melanoma | | Non-melanoma | |
|---|---|---|---|---|---|---|---|
| **Exposure** | **n/category[a]** | **Cases (% of category)** | **HR (95% CI)[b]** | **Cases (% of category)** | **HR (95% CI)[b]** | **Cases (% of category)** | **HR (95% CI)[b]** |
| **Tattooed** | | | | | | | |
| **N Total** | 105856 | 1738 (1.6%) | | 675 (0.6%) | | 1063 (1.0%) | |
| No | 98405 | 1702 (1.7%) | ref | 659 (0.7%) | ref | 1043 (1.1%) | ref |
| Yes | 7451 | 36 (0.5%) | 0.80 (0.57-1.11) | 16 (0.2%) | 0.83 (0.50-1.37) | 20 (0.3%) | 0.77 (0.49-1.20) |
| **Tattooed body surface** | | | | | | | |
| **N Total** | 105855 | 1738 (1.6%) | | 675 (0.6%) | | 1063 (1.0%) | |
| Not tattooed | 98405 | 1702 (1.7%) | ref | 659 (0.7%) | ref | 1043 (1.1%) | ref |
| 0-1 hand palm | 3867 | 28 (0.7%) | 1.00 (0.69-1.46) | 14 (0.4%) | 1.20 (0.70-2.04) | 14 (0.4%) | 0.86 (0.50-1.45) |
| 1-2 hand palms | 1978 | 6 (0.3%) | 0.61 (0.27-1.36) | 2 (0.1%) | 0.46 (0.12-1.86) | 4 (0.2%) | 0.72 (0.27-1.93) |
| >2 hand palms | 1605 | 2 (0.1%) | 0.27 (0.07-1.08) | 0 (0.0%) | No obs | 2 (0.1%) | 0.49 (0.12-1.95) |
| ***Only amongst the tattooed :*** | | | | | | | |
| **Incremental among tattooed** | 7450 | 36 (0.5%) | 0.90 (0.78-1.03) | 16 (0.2%) | 0.71 (0.47-1.07) | 20 (0.3%) | 0.95 (0.84-1.08) |
| **Sun exposure of tattoo** | | | | | | | |
| **N Total** | 5938 | 26 (0.4%) | | 12 (0.2%) | | 14 (0.2%) | |
| Not sun exposed | 1017 | 9 (0.9%) | 1.87 (0.76-4.59) | 4 (0.4%) | 3.00 (0.78-11.59) | 5 (0.5%) | 1.33 (0.36-4.94) |
| Sun exposed | 4921 | 17 (0.3%) | ref | 8 (0.2%) | ref | 9 (0.2%) | ref |

[a] Due to the different numbers of missing values for each tattoo–related variable, the number of participants (i.e. N Total for each tattoo exposure) were slightly different.

[b] Considering tattooed or not as time-varying, the dataset was split on date of first tattoo. Cox regression models were performed based on an age-time scale, with stratification of age at first exposure assessment in 2020/21 (<40, ≥40) and adjustment for biological sex, education level, smoking status, BMI, household disposable income, barriers to health care, maximum intensity of tan, skin type, frequency of sunburns during adolescence.

**Figure S1.** Directed acyclic graph (DAG) demonstrating covariate selection to estimate the association of tattoo exposure and skin cancer (including overall skin cancer, melanoma, and non-melanoma skin cancer). Tattoo is exposure and skin cancer is the outcome. Red circles are ancestor of exposure and outcome, while blue circles are ancestor of outcome.

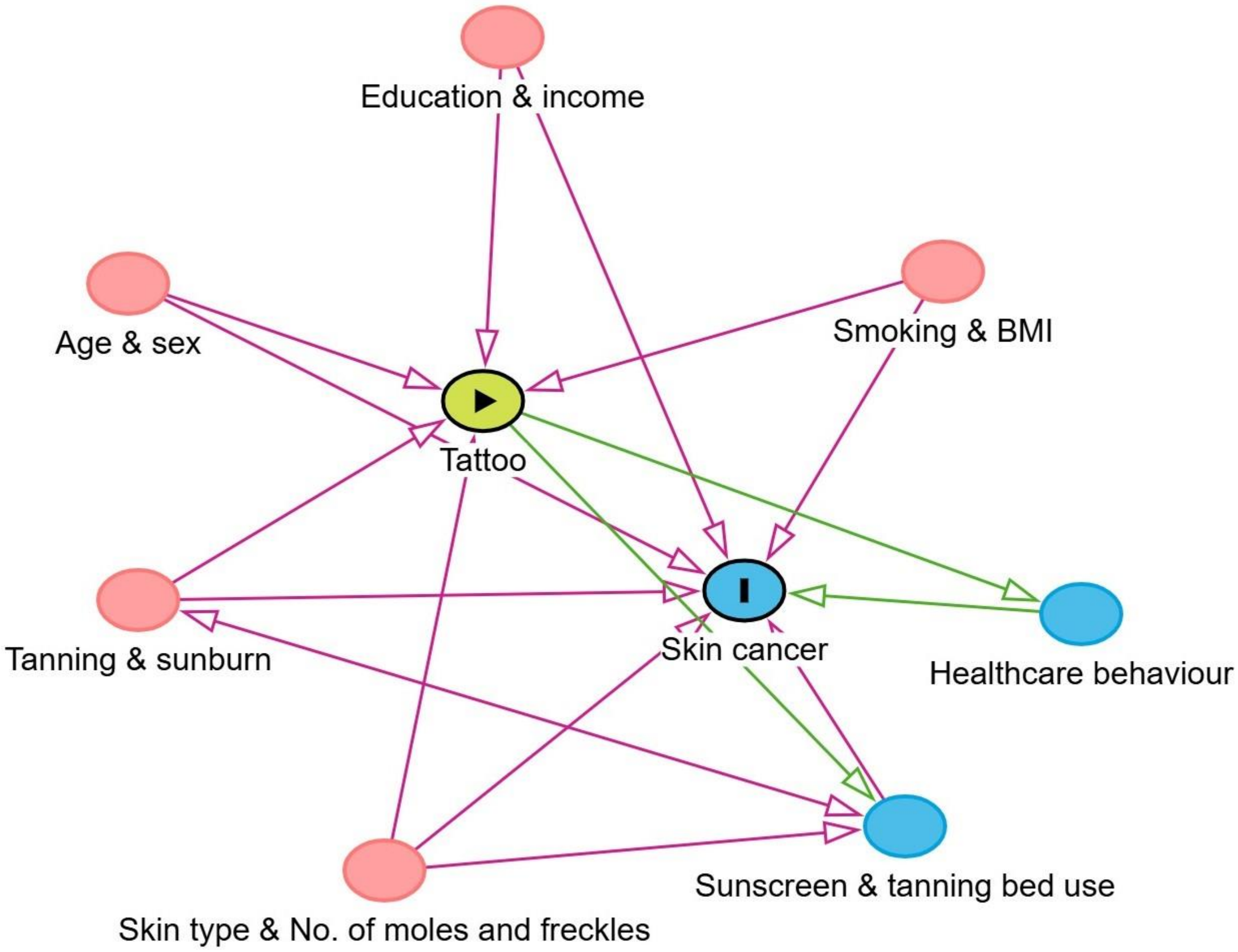
